# Blood transcriptional biomarkers of acute viral infection for detection of pre-symptomatic SARS-CoV-2 infection

**DOI:** 10.1101/2021.01.18.21250044

**Authors:** Rishi K. Gupta, Joshua Rosenheim, Lucy C. Bell, Aneesh Chandran, Jose A. Guerra-Assuncao, Gabriele Pollara, Matthew Whelan, Jessica Artico, George Joy, Hibba Kurdi, Daniel M. Altmann, Rosemary Boyton, Mala Maini, Aine McKnight, Jonathan Lambourne, Teresa Cutino-moguel, Charlotte Manisty, Thomas A. Treibel, James C Moon, Benjamin Chain, Mahdad Noursadeghi, on behalf of the COVIDsortium Investigators

**Affiliations:** Institute of Global Health, University College London, London, UK; Division of Infection and Immunity, University College London, London, UK; Barts Heart Centre, St Bartholomew’s Hospital, Barts Health NHS Trust, London, UK, London, UK; Department of Immunology and Inflammation, Imperial College London, London, UK; Lung Division, Royal Brompton & Harefield NHS Foundation Trust, London, UK; Blizard Institute, Barts and the London School of Medicine and Dentistry, Queen Mary University of London, London, UK; Department of Infection, Barts Health NHS Trust, London, UK; Department of Virology, Barts Health NHS Trust, London, UK; Institute of Cardiovascular Sciences, University College London, London, UK

## Abstract

We hypothesised that host-response biomarkers of viral infections may contribute to early identification of SARS-CoV-2 infected individuals, critical to breaking chains of transmission. We identified 20 candidate blood transcriptomic signatures of viral infection by systematic review and evaluated their ability to detect SARS-CoV-2 infection, compared to the gold-standard of virus PCR tests, among a prospective cohort of 400 hospital staff subjected to weekly testing when fit to attend work. The transcriptional signatures had limited overlap, but were mostly co-correlated as components of type 1 interferon responses. We reconstructed each signature score in blood RNA sequencing data from 41 individuals over sequential weeks spanning a first positive SARS-CoV-2 PCR, and after 6-month convalescence. A single blood transcript for IFI27 provided the highest accuracy for discriminating individuals at the time of their first positive viral PCR result from uninfected controls, with area under the receiver operating characteristic curve (AUROC) of 0.95 (95% confidence interval 0.91–0.99), sensitivity 0.84 (0.7–0.93) and specificity 0.95 (0.85–0.98) at a predefined test threshold. The test performed equally well in individuals with and without symptoms, correlated with viral load, and identified incident infections one week before the first positive viral PCR with sensitivity 0.4 (0.17–0.69) and specificity 0.95 (0.85–0.98). Our findings strongly support further urgent evaluation and development of blood IFI27 transcripts as a biomarker for early phase SARS-CoV-2 infection, for screening individuals such as contacts of index cases, in order to facilitate early case isolation and early antiviral treatments as they emerge.

## Introduction

Rapid and accurate testing is central to effective public health responses to COVID-19. Infectivity, measured by upper respiratory tract SARS-CoV-2 titres, peaks during the first week of illness *(1)*. Early case detection followed by rapid isolation of index cases, alongside contact tracing and quarantine, are therefore key interventions to interrupt onward transmission. As a proportion of individuals with SARS-CoV-2 shed virus while asymptomatic or pauci-symptomatic *(2, 3)*, there is also global interest in screening tests for at-risk individuals who do not fulfil case definition criteria and in mass testing for early case detection among the general population *(4)*.

Effective screening tests must be accurate and reliable *(5)*. However, current tools such as lateral flow assays (LFAs) for SARS-CoV-2 antigens appear to have inadequate sensitivity to effectively rule out active infection and may have limited value for contact and general population screening *(6)*. Reverse-transcriptase PCR (RT-PCR) tests, which identify viral RNA, are the current gold standard for diagnosis of SARS-CoV-2 infection but pose different challenges including test speed and the requirement of a skilled laboratory operator *(7)*. LAMP (loop-mediated isothermal amplification) assays improve on RT-PCR timings but with an associated reduction in sensitivity *(8)*. All current viral detection tests rely on swabbing of nasopharyngeal and/or oropharyngeal mucosa, the effectiveness of which is operator-dependent and prone to sampling variability. While positive SARS-CoV-2 test results are useful in clinical management and infection control settings, all available tests have false negative rates which impact on interpretation, particularly in the context of high pre-test probability in high transmission settings *(9)*.

There is therefore a clear need to expand the portfolio of tests available for identification of SARS-CoV-2 infection, for both screening and diagnostic purposes. Measurement of the host response, as opposed to viral targets, is one potential diagnostic strategy. Numerous studies have demonstrated whole-blood transcriptional perturbation during other acute viral infections *(10–13)*. A range of blood transcriptomic signatures have therefore been proposed as candidate diagnostic biomarkers for purposes including discrimination of viral from bacterial infection or no infection *(10–21)*, diagnosis of pre-symptomatic viral infection in known contacts *(22)*, diagnosis of specific viral infections *(23, 24)*, or prognostication of severity *(25)*. These signatures have not yet been evaluated for early diagnosis of pre-symptomatic or mild SARS-CoV-2 infection. We present a systematic evaluation of the potential for existing candidate whole-blood transcriptomic signatures of viral infection to predict nasopharyngeal SARS-CoV-2 PCR positivity in healthcare workers undergoing weekly testing with paired blood RNA sampling.

## Results

### Overview of study cohort

We included a total of 169 blood RNA samples from 96 participants in a nested case-control study (Fig. S1) derived from an observational healthcare worker cohort *(26–28)*. Of these, 114 samples (including 16 convalescent samples collected 6 months after infection) were obtained from 41 incident cases with SARS-CoV-2 infection, and 55 samples from uninfected controls. Participant baseline characteristics are shown in Table S1. 32/41 individuals with incident virus PCR positive infection denied any disease defining symptoms at the time of their positive PCR test, whilst 9/41 described one or more of cough, fever, or anosmia. A further 22 individuals developed symptoms during subsequent follow up.

### Overview of candidate RNA signatures for viral infection

Our systematic literature search identified 1150 titles and abstracts; 61 studies were shortlisted for full-text review. A total of 18 studies, describing 20 distinct transcriptional signatures for viral infection, met the eligibility criteria for inclusion in the final analysis (Table 1 and Fig. S2). Signatures comprised between 1 and 48 component genes and were discovered in a range of populations including children and adults with acute viral infections, and adults experimentally challenged with viruses including influenza, respiratory syncytial virus (RSV) and rhinovirus. The majority of signatures (12/20) were discovered with the objective of discriminating viral infection from bacterial or other inflammatory presentations *(10–21)*. Three aimed to discriminate viral infection from healthy individuals *(29, 30)* and two were discovered with a specific objective of diagnosing influenza infection *(23, 24)*. One signature aimed to predict the severity of RSV infection in children *(25)*. One study evaluated a pre-existing signature with the objective of identifying pre-symptomatic viral infection in individuals who were close contacts of index cases with acute viral respiratory tract infections *(22)*.

**Table 1.** Characteristics of whole-blood RNA signatures for viral infection included in analysis. Signatures are referred to by combining the first author’s name of the corresponding publication as a prefix, with number of constituent genes as a suffix. Log_2_-transformed transcripts per million data used to calculate all signatures. *Study by McClain *et al*. sought to validate a 36-transcript signature for detection of respiratory viral infections. Model coefficients for the 36-transcript model are not provided; we therefore included the two best performing single transcripts from the study in the current analysis, since they demonstrated similar performance to the full model in the original publication. ^&^Where applicable, the name of the signature from the original publication is indicated in brackets. †Defined as the sum of downregulated genes subtracted from the sum of upregulated genes. §Logistic and probit regression models were calculated on the linear predictor scale using model coefficients from original publications. Abbreviations: RSV, respiratory syncytial virus. PAM, prediction analysis of microarrays. SVM, support vector machine. LASSO, Least Absolute Shrinkage Selector Operator. RT-LAMP, Reverse Transcription Loop-mediated Isothermal Amplification. LCMV, lymphocytic choriomeningitis virus.

| Signature(s) | Model<sup>a</sup> | Discovery population(s) | Discovery setting(s) | Discovery approach | Validation population(s) | Intended application |
| --- | --- | --- | --- | --- | --- | --- |
| **AndresTerres11 (23)** | Geometric mean of all genes ("influenza meta-signature") | Five cohorts -children and adults with influenza; adults challenged with influenza; adults with bacterial pneumonia | UK, USA and Australia | Differential expression followed by leave-one-cohort-out strategy and filtering for heterogeneity of effect size, using genome-wide data | Eight cohorts - children or adults with influenza or bacterial infection; adults challenged with influenza; adults vaccinated against influenza | Influenza versus bacterial or other viral infection |
| **Henrickson16 (24)** | Difference in geometric means between upregulated and downregulated genes ("Influenza paediatric signature score") | Four cohorts -children with influenza-like illness | USA | Meta-analysis and leave-one-out strategy to identify common genes using genome-wide data | Two cohorts – children or adults with influenza. | Influenza infection versus healthy |
| **Herberg2 (10)** | Disease risk score† | Children with viral or bacterial infection | UK, USA and Spain | Elastic net followed by forward selection–partial least squares, using significantly differentially expressed transcripts | Children with bacterial or viral infection, inflammatory disease, or indeterminate diagnosis | Viral versus bacterial infection in febrile children |
| **IFI44L (14)** | N/A | Children with viral or bacterial infection (10) | UK, USA and Spain | Elastic net followed by forward selection–partial least squares, using significantly differentially expressed transcripts | Children with bacterial or viral infection | Viral versus bacterial infection in febrile children |
| **IFIT3; RSAD2\* (22)** | N/A | Three cohorts of adults challenged with rhinovirus, influenza or RSV (42) | UK and USA | Sparse latent factor regression analysis on genome-wide data (42) followed by regularised logistic regression on the resulting 30-gene signature | Close contacts of students with acute upper respiratory viral infections | Pre-symptomatic viral infection versus healthy |
| **Lopez7 (15)** | Sum of weighted gene expression values ("Bacterial versus viral classifier") | Children and adults with viral, bacterial or non-infectious acute respiratory illness (19) | USA | SVM using genome-wide data | Children with acute viral or bacterial infections (43) | Viral versus bacterial respiratory infection |
| **Lydon15 (11)** | Logistic regression ("Viral classifier")§ | Adolescents and adults with viral, bacterial or non-infectious acute respiratory illness | USA | LASSO using 87 selected target genes from previously derived signatures (19, 21) | Patients with viral/bacterial co-infection or suspected bacterial infection | Viral versus bacterial respiratory infection |
| **MX1 (29)** | N/A | N/A | N/A | Pre-selected due to biological plausibility | Adults challenged with the live yellow fever virus vaccine | Viral infection versus healthy |
| **OLFM4 (25)** | N/A | Children with RSV infection | The Netherlands | Differential expression and PAM classifier training using genome-wide data | A second cohort of children with RSV infection | Severity of RSV infection in children |
| **Pennisi2 (20)** | Disease risk score† | Children with viral or bacterial infection (10) | UK, USA and Spain | Elastic net followed by forward selection–partial least squares, using significantly differentially expressed transcripts (10), followed by selection of an adequately expressed transcript for use in RT-LAMP | Children with bacterial or viral infection | Viral versus bacterial infection in children |
| **Sampson10 (13)** | Disease risk score ("Combined SeptiCyt score") | Eight cohorts of neonates, children and adults with bacterial infections | UK, USA, Estonia and Australia | Regression analysis of transcript pairs using the 6000 most highly expressed genes from each dataset | Unselected consecutive patients presenting to the emergency department with febrile illness | Viral versus bacterial in febrile patients |
| **Sampson4 (16)** | Disease risk score ("Septicyte VIRUS") | Ten cohorts - children and adults with viral infections. Two cohorts - adults challenged with influenza. Two cohorts - macaques challenged with Lassa or LCMV. | USA, Finland, Brazil, and Australia | Regression analysis of transcript pairs using the 6000 most highly expressed genes from each dataset | Seven human cohorts and six non-human mammal cohorts, infected or challenged with viruses across all seven of the Baltimore virus classification groups | Viral versus non-viral conditions |
| **Sweeney11 (17)** | Difference in geometric means between upregulated and downregulated genes, multiplied by ratio of counts of positive to negative genes ("Sepsis metascore") | Nine cohorts of patients with sepsis or trauma | USA, Australia, Europe, Scandinavia, Canada, UK | Greedy forward search of 82 differentially expressed genes identified by multicohort analysis | Twelve cohorts of adults with viral or bacterial sepsis, or trauma | Viral or bacterial sepsis versus sterile inflammation |
| **Sweeney7 (12)** | Difference in geometric means between upregulated and downregulated genes, multiplied by ratio of counts of positive to negative genes ("Bacterial/viral metascore") | Eight cohorts of children and adults with viral and bacterial infections | USA, Australia, UK | Greedy forward search of 72 differentially expressed genes identified by multicohort analysis | Twenty-four cohorts of children and adults with viral or bacterial infections, or healthy controls | Viral versus bacterial infection |
| **Trouillet-Assant6 (18)** | Median expression of 6 interferon-stimulated genes ("Interferon score (44)") | N/A | N/A | Differential expression using 15 preselected interferon-stimulated genes | Febrile children with bacterial or viral infection | Viral versus bacterial infection in febrile children |
| **Tsalik33 (19)** | Logistic regression ("Viral ARI classifier")§ | Children and adults with viral, bacterial or non-infectious acute respiratory illness, and healthy controls | USA | LASSO using the 40% of microarray probes with the largest variance after batch correction | Five cohorts – children or adults with viral, bacterial or non-infectious respiratory illness, or viral/bacterial co-infection | Viral versus bacterial acute respiratory illness |
| **Yu3; IFI27 (30)** | 1. Mean expression ("non-RSV infections vs. controls")¶¶¶<br>2. N/A | Children with acute respiratory illness and a positive result for a viral infection on a nasopharyngeal swab | USA | Modified supervised principal component analysis using all expressed transcripts | Children with RSV or rhinovirus infection | Viral versus healthy in children |
| **Zaas48 (21)** | Probit regression ("Viral classifier")§ | Two cohorts of adults challenged with influenza A H3N2 or H1N1 | USA | Elastic net using 48 selected genes comprised of 29 derived as a signature in a previous study (42), 7 shown to be downregulated in analysis of influenza challenge time course data (45) and 12 control genes | Adults presenting to the emergency department with fever and healthy controls | Viral versus bacterial acute respiratory illness |

In most cases there was little overlap between the constituent genes in each signature, but most signatures showed moderate to strong correlation, which was only partly explained by overlapping constituent genes (Fig. 1A–C). Bioinformatic analysis of the integrated list of constituent genes to identify upstream regulators using Ingenuity Pathway Analysis was consistent with type 1 interferon regulation of these genes to explain the strong correlation between signatures despite limited overlap of their constituents.

**Fig. 1.**
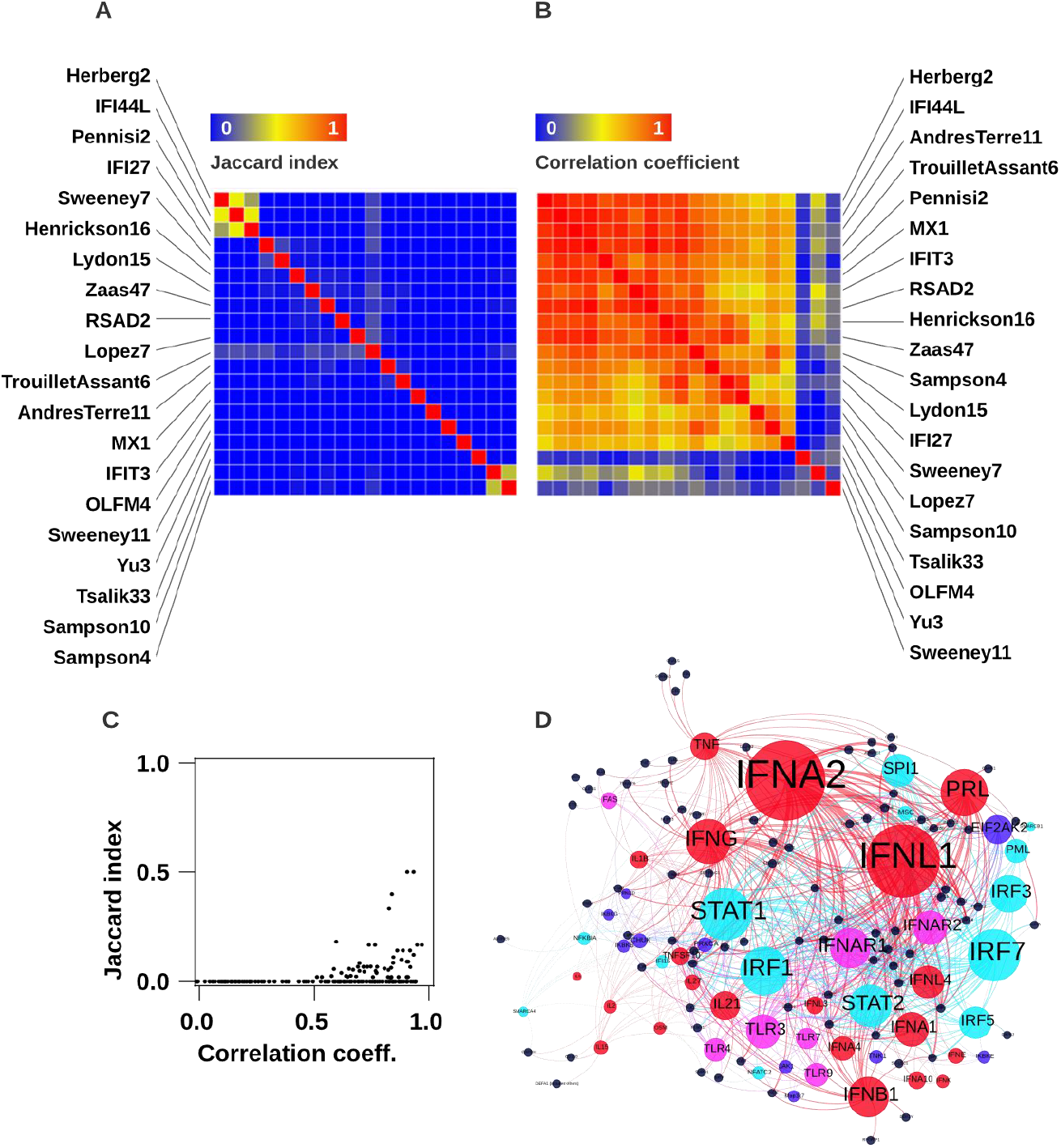
Correlation and Jaccard indices for all RNA signatures for viral infection included in analysis. **(A**) Jaccard index intersect of constituent genes for all pairs of signatures clustered by Euclidean distance. **(B)** Spearman rank correlation coefficients for all pairs of signatures clustered by 1-Spearmean rank distance **(C)** relationship between pairwise Jaccard indices and Spearman rank correlation coefficients. **(D)** Network plot of significantly enriched predicted upstream regulators by cytokine (red nodes), transmembrane receptors (purple nodes), kinase (dark blue nodes) and transcription factors (light blue nodes) of all constituent genes in any signature (black nodes). Size of upstream regulator nodes proportional to statistical enrichment (-log_10_ FDR).

### Diagnostic accuracy of RNA signatures for SARS-CoV-2 infection

Among all the signatures, the transcript for Interferon Alpha Inducible Protein 27 (IFI27) by itself provided the best discrimination of contemporaneous SARS-CoV-2 infection by nasopharyngeal PCR, compared to uninfected controls, achieving an AUROC of 0.95 (95% confidence interval 0.91–0.99). Using a pre-specified Z2 cut-off based on two standard deviations above the mean of the uninfected control samples, IFI27 had a sensitivity of 0.84 (0.70–0.93) and specificity of 0.95 (0.85–0.98). Three other candidate signatures (Sweeney7, Zaas48 and Pennisi2) had statistically equivalent accuracy to IFI27 using paired DeLong tests (Table 2). Fig. S3 shows constituent genes for these four best performing signatures; only one of these (Pennisi2) did not include IFI27. Exclusion of participants with contemporaneous case-defining symptoms at the time of SARS-CoV-2 infection (n=9) had no significant impact on the primary analysis (Table S2). Scores for each of the four best performing signatures were inversely correlated with SARS-CoV-2 RT-PCR cycle thresholds, also independent of symptoms, suggesting that higher viral loads were associated with higher signature scores (Figure 2B; Spearman rank correlation coefficients −0.61 to −0.69).

**Table 2.** Validation metrics of whole-blood RNA signatures for discrimination of participants with PCR-confirmed SARS-CoV-2 infection at first week of PCR-positivity (PCR+ve_0). Discrimination is shown as area under the receiver operating characteristic curve (AUROC). Sensitivity and specificity are shown using pre-defined thresholds of 2 standard deviations above the mean of the uninfected control population (Z2). P values show pairwise comparisons to best performing signature with Benjamini-Hochberg adjustment (false discovery rate 0.05). All metrics as shown as point estimates (95% confidence intervals). Equivalent table for discrimination of participants with SARS-CoV-2 infection one week prior to PCR-positivity (PCR+ve_-1), compared to uninfected controls, is shown in Table S3.

| Signature | AUROC | Sensitivity | Specificity | p |
| --- | --- | --- | --- | --- |
| IFI27 | 0.95 (0.91 - 0.99) | 0.84 (0.7 - 0.93) | 0.95 (0.85 - 0.98) | NA |
| Sweeney7 | 0.95 (0.91 - 0.99) | 0.82 (0.67 - 0.91) | 0.95 (0.85 - 0.98) | 0.851 |
| Zaas48 | 0.93 (0.88 - 0.98) | 0.61 (0.45 - 0.74) | 0.95 (0.85 - 0.98) | 0.088 |
| Pennisi2 | 0.91 (0.86 - 0.96) | 0.58 (0.42 - 0.72) | 0.95 (0.85 - 0.98) | 0.088 |
| IFI44L | 0.9 (0.84 - 0.96) | 0.55 (0.4 - 0.7) | 0.95 (0.85 - 0.98) | 0.039 |
| AndresTerre11 | 0.89 (0.83 - 0.95) | 0.55 (0.4 - 0.7) | 0.95 (0.85 - 0.98) | 0.021 |
| Henrickson16 | 0.89 (0.82 - 0.96) | 0.55 (0.4 - 0.7) | 0.93 (0.83 - 0.97) | 0.009 |
| TrouilletAssant6 | 0.87 (0.8 - 0.94) | 0.53 (0.37 - 0.68) | 0.93 (0.83 - 0.97) | 0.008 |
| Lydon15 | 0.86 (0.79 - 0.94) | 0.58 (0.42 - 0.72) | 0.95 (0.85 - 0.98) | 0.005 |
| Herberg2 | 0.84 (0.76 - 0.92) | 0.5 (0.35 - 0.65) | 0.93 (0.83 - 0.97) | 0.003 |
| Sampson4 | 0.84 (0.76 - 0.92) | 0.5 (0.35 - 0.65) | 0.93 (0.83 - 0.97) | 0.003 |
| Sampson10 | 0.83 (0.74 - 0.92) | 0.5 (0.35 - 0.65) | 0.95 (0.85 - 0.98) | 0.002 |
| RSAD2 | 0.83 (0.74 - 0.91) | 0.47 (0.32 - 0.63) | 0.93 (0.83 - 0.97) | 0.002 |
| MX1 | 0.82 (0.74 - 0.91) | 0.45 (0.3 - 0.6) | 0.95 (0.85 - 0.98) | 0.002 |
| Tsalik33 | 0.79 (0.7 - 0.89) | 0.39 (0.26 - 0.55) | 0.98 (0.9 - 1) | 0.001 |
| Lopez7 | 0.79 (0.69 - 0.88) | 0.37 (0.23 - 0.53) | 0.98 (0.9 - 1) | 0.001 |
| IFIT3 | 0.75 (0.64 - 0.86) | 0.45 (0.3 - 0.6) | 0.93 (0.83 - 0.97) | 0.000 |
| OLFM4 | 0.62 (0.51 - 0.74) | 0.03 (0 - 0.13) | 0.98 (0.9 - 1) | 0.000 |
| Sweeney11 | 0.6 (0.48 - 0.73) | 0.16 (0.07 - 0.3) | 0.96 (0.88 - 0.99) | 0.000 |
| Yu3 | 0.59 (0.47 - 0.71) | 0.05 (0.01 - 0.17) | 1 (0.93 - 1) | 0.000 |

**Figure 2.**
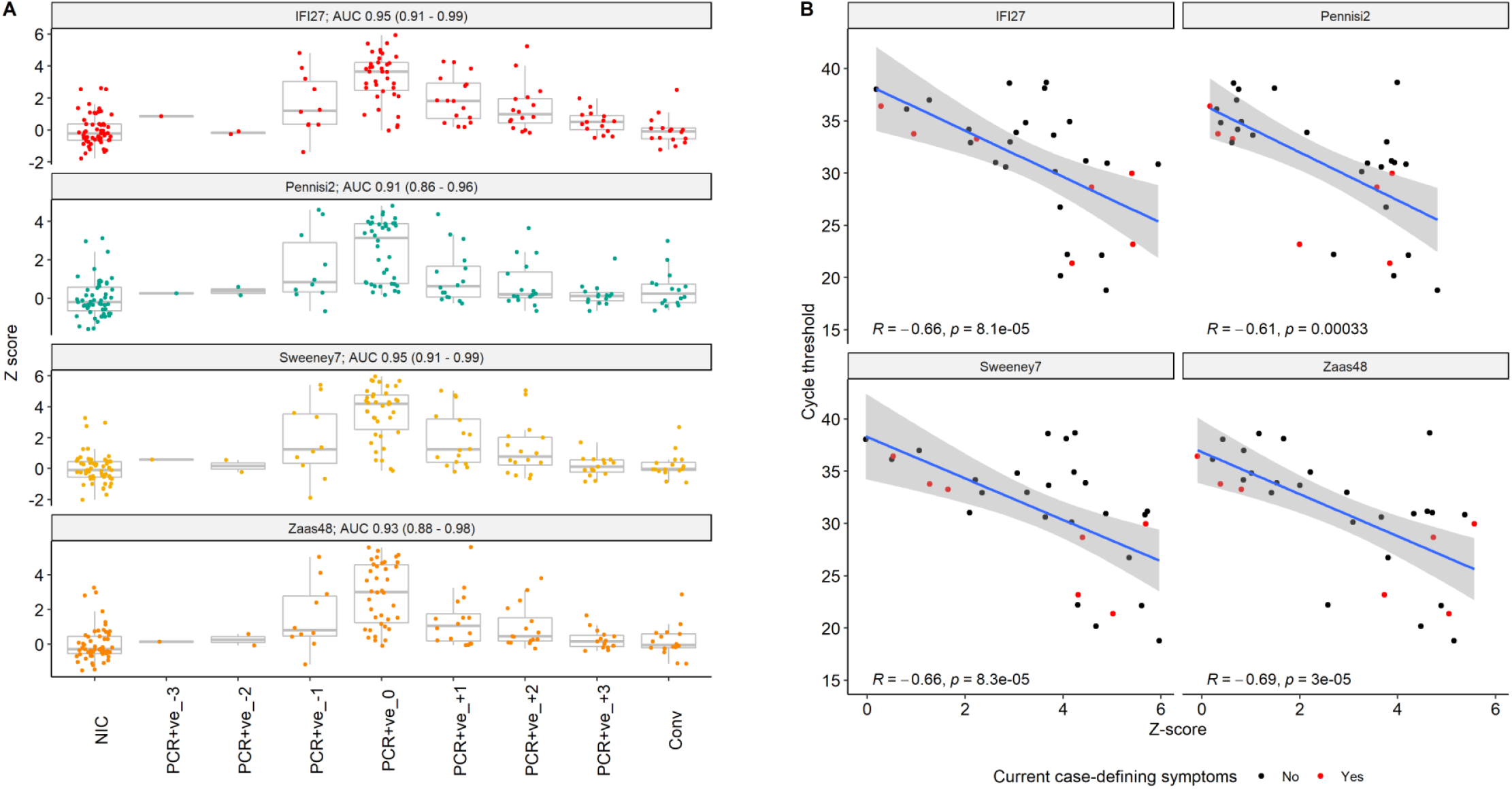
Four best performing RNA signatures for discriminating participants with contemporaneous PCR-confirmed SARS-CoV-2 infection, compared to uninfected controls. Shown as **(A)** Z scores stratified by interval to positive SARS-CoV-2 PCR; and **(B)** Z scores vs. contemporaneous PCR cycle threshold for SARS-CoV-2 ORF1. NIC = non-infected controls; Conv = convalescent samples, collected at study week 24. AUC = area under the receiver operating characteristic curve (95% CI) for discriminating participants with contemporaneous PCR-confirmed SARS-CoV-2 infection (PCR+ve_0 group), compared to uninfected controls. R = Spearman rank correlation coefficient.

Longitudinal expression of the four best performing signatures peaked at the week of first virus PCR-positivity and had normalised at the point of convalescent sampling (Fig. 2). Importantly, however, measurements in the week preceding the first positive virus PCR were higher than uninfected controls and convalescent samples. AUROCs for discrimination between samples taken in the week prior to first SARS-CoV-2 detection and uninfected controls revealed statistically significant discrimination, but were lower than those for contemporaneous virus PCR positivity (Table S3). For illustration, IFI27 predicted infection one week before a positive virus PCR test with an AUROC of 0.79 (0.6–0.98). At a Z2 cut-off this achieved sensitivity of 0.4 (0.17–0.69) and specificity of 0.95 (0.85–0.98).

## Discussion

To our knowledge, this is the first evaluation of host transcriptomic signatures for detection of pre-symptomatic SARS-CoV-2 infection. Using a longitudinal blood transcriptomic dataset prospectively collected from London healthcare workers during the first wave of the COVID-19 pandemic, we systematically compared the diagnostic accuracy of 20 candidate transcriptional signatures originally discovered in a wide range of viral infection cohorts. We found that four candidate signatures had high accuracy (AUROCs 0.91–0.95) for discriminating individuals with acute contemporaneous SARS-CoV-2 infection from uninfected controls. Three of the four signatures contained the interferon-stimulated gene *IFI27*, which was by itself the top-performing biomarker, originally discovered in a paediatric cohort *(30)* to discriminate RSV infections from controls.

The candidate signatures evaluated in the present study are collectively associated with type 1 IFN responses, which are a canonical feature of antiviral host defences. The importance of this response in SARS-CoV-2 infection is highlighted by the association of severe COVID-19 with both of loss-of-function genetic variation in various components of type 1 IFN pathways and with anti-type 1 IFN antibodies *(31, 32)*. IFI27 is best characterised for its functional role in type 1 IFN mediated apoptosis as a component of anti-tumour effects of IFNs *(33)*. Differential regulation of IFN inducible genes might explain why expression of IFI27 transcripts outperforms other type 1 IFN signatures and merits investigation in future work to evaluate its significance in the anti-viral response.

A key feature of our study is that all participants self-declared as fit to work when attending study visits, including at the time of their first positive SARS-CoV-2 PCR test, when most were asymptomatic. We also found detectable expression of the signatures in blood transcriptomes collected at the study visit one week prior to virus PCR test positivity among a subset of participants. Our data therefore demonstrate that measurable type 1 IFN-stimulated responses to SARS-CoV-2 precede the onset of symptoms, and in some individuals may predate detectable viral RNA on RT-PCR testing. We propose that novel diagnostic tests that detect transcripts (or associated protein targets) from the top-performing candidate signatures could be valuable tools in the rapid detection and isolation of individuals in the very earliest stages of pre-clinical infection. Importantly, these signatures also correlated with viral load independently of symptoms, indicating that they have strong potential to identify the most infectious individuals, critical to breaking chains of transmission.

A key strength of our study was the weekly longitudinal follow-up of study participants which enabled detailed characterisation of the study cohort, including contemporaneous capture of blood RNA samples at the point of SARS-CoV-2 PCR positivity in pre-symptomatic and asymptomatic infection. In addition, we performed a comprehensive systematic literature search to identify candidate blood transcriptional signatures for viral infection. This enabled direct head-to-head assessments of their diagnostic accuracy for SARS-CoV-2 infection and will provide a framework for future systematic evaluations of blood transcriptional biomarkers for viral infections. Our findings focus on early pre-symptomatic infection and may not be generalisable in moderate to severe COVID-19. In addition, we have not sought to evaluate discrimination between SARS-CoV-2 and other acute viral infections. In view of their discovery in a range of viral infections, we expect these signatures to serve as non-specific biomarkers of acute viral infection. Nonetheless, their sensitivity for detecting pre-symptomatic infection offers potential clinical utility for screening contacts of index cases of SARS-CoV-2 in order to inform infection control management, and stratify the need for confirmatory viral PCR testing.

In summary, we have shown that a single transcript (IFI27) detects SARS-CoV-2 infection with high accuracy. If translated to a near-patient diagnostic test *(34, 35)*, this transcript could have significant clinical utility by facilitating early case detection.

## Materials and Methods

### Ethical approval

The study was approved by a UK Research Ethics Committee (South Central - Oxford A Research Ethics Committee, reference 20/SC/0149). All participants provided written informed consent.

### Study design

We undertook a case-control study nested within our COVIDsortium health care worker cohort. Participant screening, study design, sample collection, and sample processing have been described in detail previously *(26–28)* and the study is registered at ClinicalTrials.gov (NCT04318314). Briefly, healthcare workers were recruited at St Bartholomew’s Hospital, London, UK in the week of lockdown in the United Kingdom (between 23^rd^ and 31^st^ March 2020). Participants underwent weekly evaluation using a questionnaire and biological sample collection (including serological assays) for up to 16 weeks when fit to attend work at each visit, with further follow up samples collected at 6 months.

Participants with available blood RNA samples who had PCR-confirmed SARS-CoV-2 infection (Roche cobas® diagnostic test platform) at any time point were included as ‘cases’. A subset of consecutively recruited participants without evidence of SARS-CoV-2 infection on nasopharyngeal swabs and who remained seronegative by both Euroimmun anti-S1 spike protein and Roche anti-nucleocapsid protein throughout follow-up were included as uninfected controls.

### Systematic search for candidate transcriptional signatures

We performed a systematic literature search of peer-reviewed publications in order to identify concise blood transcriptional signatures discovered or applied with a primary objective of diagnosis or assessment of severity of viral infection from blood or PBMC samples. We searched Medline on 12/10/2020 using comprehensive MeSH and key word terms for “viral infection”, “transcriptome”, “biomarker” and “blood” (full search strategy shown in Table S4). Additional studies were identified in reference lists and from expert consultation. Titles and abstracts were initially screened by two independent reviewers; full-text review was performed for shortlisted articles to determine eligibility and conflicts were resolved through discussion and arbitration by a third reviewer where required. We focused on ‘concise’ signatures that might be more amenable to translation to diagnostic tests and defined this as any signature discovered using a defined approach to feature selection to reduce the number of constituent genes, as previously *(36)*. We required that gene names that comprised the signature were publicly available, along with the corresponding signature equation or model coefficients, and that the signature must be validated in at least one independent test or validation set in order to prioritise signatures discovered from higher quality studies. Where multiple signatures were discovered for the same intended purpose and from the same discovery cohort, we included the signature with highest discrimination (as defined by the AUROC) in the validation data, or the signature with fewest number of genes where accuracy was equivalent.

For each signature that met our eligibility criteria, we extracted constituent genes, modelling approaches and coefficients to enable independent reconstruction of signature scores. Extraction was performed by a single reviewer and was verified by a second reviewer.

### Blood RNA sequencing

For ‘cases’, we included all available RNA samples, including convalescent samples at week 24 of follow-up for a subset of participants. For uninfected controls, we included baseline samples only. Genome wide mRNA sequencing was performed as previously described *(37)*, resulting in a median of 26 million (range, 19.8–32.4 million) 41 bp paired-end reads per sample. RNAseq data were mapped to the reference transcriptome (Ensembl Human GRCh38 release 100) using Kallisto *(38)*. The transcript-level output counts and transcripts per million (TPM) values were summed on gene level and annotated with Ensembl gene ID, gene name, and gene biotype using the R/Bioconductor packages tximport and BioMart *(39, 40)*.

### Data analysis

For each eligible signature, we reconstructed signature scores as per the original authors’ descriptions. For logistic and probit regression models, we calculated scores on the linear predictor scale by summing the expression of each constituent gene multiplied by its coefficient. Scores for each signature were standardised to Z scores using the mean and standard deviation among the uninfected control population. Scores that were designed to decrease in viral infection were multiplied by −1 in order to ensure that higher scores should be associated with higher risk of viral infection across all candidate signatures.

The primary outcome was the AUROC for discriminating participants with PCR-confirmed SARS-CoV-2 infection during their first week of PCR positivity, from uninfected control samples. The secondary outcome was the AUROC for discriminating participants with PCR-confirmed SARS-CoV-2 infections in the week prior to first PCR positivity, from uninfected control samples. We calculated corresponding sensitivities and specificities for each signature for the primary and secondary outcomes using pre-specified cut-offs based on two standard deviations above the mean of the uninfected controls, as previously *(36)*. In order to identify the subset of best performing signatures, we performed pairwise DeLong tests to the signature with the highest AUROC for the primary outcome (or most parsimonious in the event of equal performance), with adjustment for multiple testing using a Benjamini-Hochberg correction *(41)*. Signatures were considered to have statistically inferior accuracy to the best performing signature if adjusted p<0.05. We also performed a sensitivity analysis for the primary outcome, excluding participants with positive SARS-CoV-2 swabs who reported contemporaneous case-defining symptoms at the time of sampling.

Upstream analysis of transcriptional regulation of the constituent genes in the candidate signatures was performed using Ingenuity Pathway Analysis (Qiagen, Venlo, The Netherlands) and visualized as network diagrams in Gephi v0.9.2, depicting all statistically overrepresented molecules predicted to be upstream >2 target genes, as previously *(36)*. We evaluated pairwise Spearman rank and Jaccard indices between each candidate signature in order to quantify correlations and proportions of intersecting genes between signatures.

## Data Availability

Open access to RNAseq data and associated essential metadata are available under accession no E-MTAB-10022 at ArrayExpress (https://www.ebi.ac.uk/arrayexpress/).

## Role of the funding source

The funder had no role in study design, data collection, data analysis, data interpretation, writing of the report, or decision to submit for publication. The corresponding authors had full access to all the data in the study and had final responsibility for the decision to submit for publication.

## Supplementary Materials

Fig. S1. CONSORT (Consolidated Standards of Reporting Trials) flow diagram.

Fig. S2. PRISMA (Preferred Reporting Items for Systematic Reviews and Meta-Analyses) flow-chart of systematic review process.

Fig S3. Constituent genes of best-performing RNA signatures.

Table S1. Search strategy for systematic Medline search.

Table S2. Validation metrics of whole-blood RNA signatures at week of first week of PCR-positivity.

Table S3. Validation metrics of whole-blood RNA signatures at week prior to first week of PCR-positivity.

Table S4. Baseline characteristics of the study cohort.

Table S5. Full list of COVIDsortium investigators.

## Funding

Funding for COVIDsortium was donated by individuals, charitable Trusts, and corporations including Goldman Sachs, Citadel and Citadel Securities, The Guy Foundation, GW Pharmaceuticals, Kusuma Trust, and Jagclif Charitable Trust, and enabled by Barts Charity with support from UCLH Charity. RKG is funded by National Institute for Health Research (DRF-2018-11-ST2-004). JCM, CM and TAT are directly and indirectly supported by the University College London Hospitals (UCLH) and Barts NIHR Biomedical Research Centres and through the British Heart Foundation (BHF) Accelerator Award (AA/18/6/34223). TAT is funded by a BHF Intermediate Research Fellowship (FS/19/35/34374). MN is supported by the Wellcome Trust (207511/Z/17/Z) and by NIHR Biomedical Research Funding to UCL and UCLH. RJB/DMA are supported by UKRI/MRC Newton (MR/S019553/1, MR/R02622X/1 and MR/V036939/1MR/S019553/1 and MR/R02622X/1), NIHR Imperial Biomedical Research Centre (BRC):ITMAT, Cystic Fibrosis Trust SRC, and Horizon 2020 Marie Curie Actions. MKM is supported by the UKRI/NIHR UK-CIC grant, a Wellcome Trust Investigator Award (214191/Z/18/Z) and a CRUK Immunology grant (26603) AM is supported by Rosetrees Trust, The John Black Charitable Foundation, and Medical College of St Bartholomew’s Hospital Trust.

## Author contributions

CM, TAT, JCM and MN designed the study. JR, AC, MJ, GJ, MA, JL, TCM, CM and TAT acquired the data. RKG, JR, LB and MN analysed the data. RKG, LB and MN designed and performed the systematic literature search. RKG, JR, LB, DA, RB, MM, AM, BMC, CM, TAT, JCM and MN interpreted the data. RKG, JR, LB and MN wrote the manuscript with input from all the authors.

## Competing interests

The authors declare no conflict of interests.

## Data sharing statement

Applications for access to the individual participant de-identified data (including data dictionaries) and samples can be made to the access committee via an online application https://covid-consortium.com/application-for-samples/. Each application will be reviewed, with decisions to approve or reject an application for access made on the basis of (i) accordance with participant consent and alignment to the study objectives (ii) evidence for the capability of the applicant to undertake the specified research and (iii) availability of the requested samples. The use of all samples and data will be limited to the approved application for access and stipulated in the material and data transfer agreements between participating sites and investigators requesting access.

## Notes

### Competing Interest Statement

The authors have declared no competing interest.

### Clinical Trial

NCT04318314

### Clinical Protocols

https://wellcomeopenresearch.org/articles/5-179

## References and Notes

1. M. Cevik, M. Tate, O. Lloyd, A. E. Maraolo, J. Schafers, A. Ho, SARS-CoV-2, SARS-CoV, and MERS-CoV viral load dynamics, duration of viral shedding, and infectiousness: a systematic review and meta-analysis, The Lancet Microbe 2 (2020), doi:10.1016/S2666-5247(20)30172-5.

2. M. R. Kasper, J. R. Geibe, C. L. Sears, A. J. Riegodedios, T. Luse, A. M. V. Thun, M. B. McGinnis, N. Olson, D. Houskamp, R. Fenequito, T. H. Burgess, A. W. Armstrong, G. DeLong, R. J. Hawkins, B. L. Gillingham, An Outbreak of Covid-19 on an Aircraft Carrier, New England Journal of Medicine 383 (2020), doi:10.1056/NEJMoa2019375.

3. A. A. Sayampanathan, C. S. Heng, P. H. Pin, J. Pang, T. Y. Leong, V. J. Lee, Infectivity of asymptomatic versus symptomatic COVID-19, The Lancet 397 (2020), doi:10.1016/S0140-6736(20)32651-9.

4. T. Burki, Mass testing for COVID-19, The Lancet Microbe 1, e317 (2020).

5. M. J. Dobrow, V. Hagens, R. Chafe, T. Sullivan, L. Rabeneck, Consolidated principles for screening based on a systematic review and consensus process, CMAJ : Canadian Medical Association Journal 190, E422 (2018).

6. J. J. Deeks, A. E. Raffle, Lateral flow tests cannot rule out SARS-CoV-2 infection, BMJ 371 (2020), doi:10.1136/bmj.m4787.

7. N. Younes, D. W. Al-Sadeq, H. AL-Jighefee, S. Younes, O. Al-Jamal, H. I. Daas, Hadi. M. Yassine, G. K. Nasrallah, Challenges in Laboratory Diagnosis of the Novel Coronavirus SARS-CoV-2, Viruses 12 (2020), doi:10.3390/v12060582.

8. M. Mustafa Hellou, A. Górska, F. Mazzaferri, E. Cremonini, E. Gentilotti, P. De Nardo, I. Poran, M. M. Leeflang, E. Tacconelli, M. Paul, Nucleic acid amplification tests on respiratory samples for the diagnosis of coronavirus infections: a systematic review and meta-analysis, Clin Microbiol Infect (2020), doi:10.1016/j.cmi.2020.11.002.

9. J. Watson, P. F. Whiting, J. E. Brush, Interpreting a covid-19 test result, BMJ 369 (2020), doi:10.1136/bmj.m1808.

10. J. A. Herberg, M. Kaforou, V. J. Wright, H. Shailes, H. Eleftherohorinou, C. J. Hoggart, M. Cebey-López, M. J. Carter, V. A. Janes, S. Gormley, C. Shimizu, A. H. Tremoulet, A. M. Barendregt, A. Salas, J. Kanegaye, A. J. Pollard, S. N. Faust, S. Patel, T. Kuijpers, F. Martinón-Torres, J. C. Burns, L. J. M. Coin, M. Levin, IRIS Consortium, Diagnostic Test Accuracy of a 2-Transcript Host RNA Signature for Discriminating Bacterial vs Viral Infection in Febrile Children, JAMA 316, 835–845 (2016).

11. E. C. Lydon, R. Henao, T. W. Burke, M. Aydin, B. P. Nicholson, S. W. Glickman, V. G. Fowler, E. B. Quackenbush, C. B. Cairns, S. F. Kingsmore, A. K. Jaehne, E. P. Rivers, R. J. Langley, E. Petzold, E. R. Ko, M. T. McClain, G. S. Ginsburg, C. W. Woods, E. L. Tsalik, Validation of a host response test to distinguish bacterial and viral respiratory infection, EBioMedicine 48, 453–461 (2019).

12. T. E. Sweeney, H. R. Wong, P. Khatri, Robust classification of bacterial and viral infections via integrated host gene expression diagnostics, Science Translational Medicine 8, 346ra91–346ra91 (2016).

13. D. Sampson, T. D. Yager, B. Fox, L. Shallcross, L. McHugh, T. Seldon, A. Rapisarda, R. A. Hendriks, R. B. Brandon, K. Navalkar, N. Simpson, S. Stafford, E. Gil, C. Venturini, E. Tsaliki, J. Roe, B. Chain, M. Noursadeghi, Blood transcriptomic discrimination of bacterial and viral infections in the emergency department: a multi-cohort observational validation study, BMC Medicine 18, 185 (2020).

14. A. Gómez-Carballa, M. Cebey-López, J. Pardo-Seco, R. Barral-Arca, I. Rivero-Calle, S. Pischedda, M.J. Currás-Tuala, J. Gómez-Rial, F. Barros, F. Martinón-Torres, A. Salas, A qPCR expression assay of IFI44L gene differentiates viral from bacterial infections in febrile children, Sci Rep 9, 1–12 (2019).

15. R. Lopez, R. Wang, G. Seelig, A molecular multi-gene classifier for disease diagnostics, Nature Chem 10, 746–754 (2018).

16. D. L. Sampson, B. A. Fox, T. D. Yager, S. Bhide, S. Cermelli, L. C. McHugh, T. A. Seldon, R. A. Brandon, E. Sullivan, J. J. Zimmerman, M. Noursadeghi, R. B. Brandon, A Four-Biomarker Blood Signature Discriminates Systemic Inflammation Due to Viral Infection Versus Other Etiologies, Sci Rep 7, 1–17 (2017).

17. T. E. Sweeney, A. Shidham, H. R. Wong, P. Khatri, A comprehensive time-course–based multicohort analysis of sepsis and sterile inflammation reveals a robust diagnostic gene set, Science Translational Medicine 7, 287ra71–287ra71 (2015).

18. S. Trouillet-Assant, S. Viel, A. Ouziel, L. Boisselier, P. Rebaud, R. Basmaci, N. Droz, A. Belot, S. Pons, K. Brengel-Pesce, Y. Gillet, E. Javouhey, A. S. Group, M. Mommert, A. Guichard, F. Bartolo, L. Generenaz, A. Pachot, C. Capella, L. Hees, E. Mezgueldi, C. Toumi, C. Bouchiat-Sarabi, J. Casalegno, A. Portefaix, R. D. de Cambronne, M. Perret, Type I Interferon in Children with Viral or Bacterial Infections, Clin Chem 66, 802–808 (2020).

19. E. L. Tsalik, R. Henao, M. Nichols, T. Burke, E. R. Ko, M. T. McClain, L. L. Hudson, A. Mazur, D. H. Freeman, T. Veldman, R. J. Langley, E. B. Quackenbush, S. W. Glickman, C. B. Cairns, A. K. Jaehne, E. P. Rivers, R. M. Otero, A. K. Zaas, S. F. Kingsmore, J. Lucas, V. G. Fowler, L. Carin, G. S. Ginsburg, C. W. Woods, Host gene expression classifiers diagnose acute respiratory illness etiology, Science Translational Medicine 8, 322ra11–322ra11 (2016).

20. I. Pennisi, J. Rodriguez-Manzano, A. Moniri, M. Kaforou, J. A. Herberg, M. Levin, P. Georgiou, Translation of a host blood RNA Signature distinguishing bacterial from viral infection into a platform suitable for development as a point-of-care test, JAMA Pediatrics (2021), doi:10.1001/jamapediatrics.2020.5227.

21. A. K. Zaas, T. Burke, M. Chen, M. McClain, B. Nicholson, T. Veldman, E. L. Tsalik, V. Fowler, E. P. Rivers, R. Otero, S. F. Kingsmore, D. Voora, J. Lucas, A. O. Hero, L. Carin, C. W. Woods, G. S. Ginsburg, A Host-Based RT-PCR Gene Expression Signature to Identify Acute Respiratory Viral Infection, Science Translational Medicine 5, 203ra126–203ra126 (2013).

22. M. T. McClain, F. J. Constantine, B. P. Nicholson, M. Nichols, T. W. Burke, R. Henao, D. C. Jones, L. L. Hudson, L. B. Jaggers, T. Veldman, A. Mazur, L. P. Park, S. Suchindran, E. L. Tsalik, G. S. Ginsburg, C. W. Woods, A blood-based host gene expression assay for early detection of respiratory viral infection: an index-cluster prospective cohort study, The Lancet Infectious Diseases 0 (2020), doi:10.1016/S1473-3099(20)30486-2.

23. M. Andres-Terre, H. M. McGuire, Y. Pouliot, E. Bongen, T. E. Sweeney, C. M. Tato, P. Khatri, Integrated, Multi-cohort Analysis Identifies Conserved Transcriptional Signatures across Multiple Respiratory Viruses, Immunity 43, 1199 (2015).

24. S. E. Henrickson, S. Manne, D. V. Dolfi, K. D. Mansfield, K. Parkhouse, R. D. Mistry, E. R. Alpern, S. E. Hensley, K. E. Sullivan, S. E. Coffin, E. J. Wherry, Genomic Circuitry Underlying Immunological Response to Pediatric Acute Respiratory Infection, Cell Reports 22, 411–426 (2018).

25. H. K. Brand, I. M. L. Ahout, D. de Ridder, A. van Diepen, Y. Li, M. Zaalberg, A. Andeweg, N. Roeleveld, R. de Groot, A. Warris, P. W. M. Hermans, G. Ferwerda, F. J. T. Staal, Olfactomedin 4 Serves as a Marker for Disease Severity in Pediatric Respiratory Syncytial Virus (RSV) Infection, PLOS ONE 10, e0131927 (2015).

26. J. B. Augusto, K. Menacho, M. Andiapen, R. Bowles, M. Burton, S. Welch, A. N. Bhuva, A. Seraphim, C. Pade, G. Joy, M. Jensen, R. H. Davies, G. Captur, M. Fontana, H. Montgomery, B. O’Brien, A. D. Hingorani, T. Cutino-Moguel, Á. McKnight, H. Abbass, M. Alfarih, Z. Alldis, G. L. Baca, A. Boulter, O. V. Bracken, N. Bullock, N. Champion, C. Chan, X. Couto-Parada, K. Dieobi-Anene, K. Feehan, G. Figtree, M. C. Figtree, M. Finlay, N. Forooghi, J. M. Gibbons, P. Griffiths, M. Hamblin, L. Howes, I. Itua, M. Jones, V. Jardim, V. Kapil, W.-Y. J. Lee, V. Mandadapu, C. Mfuko, O. Mitchelmore, S. Palma, K. Patel, S. E. Petersen, B. Piniera, R. Raine, A. Rapala, A. Richards, G. Sambile, J. C. de Sousa, M. Sugimoto, G. D. Thornton, J. Artico, D. Zahedi, R. Parker, M. Robathan, L. M. Hickling, N. Ntusi, A. Semper, T. Brooks, J. Jones, A. Tucker, J. Veerapen, M. Vijayakumar, T. Wodehouse, L. Wynne, T. A. Treibel, M. Noursadeghi, C. Manisty, J. C. Moon, Healthcare Workers Bioresource: Study outline and baseline characteristics of a prospective healthcare worker cohort to study immune protection and pathogenesis in COVID-19, Wellcome Open Research 5, 179 (2020).

27. T. A. Treibel, C. Manisty, M. Burton, Á. McKnight, J. Lambourne, J. B. Augusto, X. Couto-Parada, T. Cutino-Moguel, M. Noursadeghi, J. C. Moon, COVID-19: PCR screening of asymptomatic health-care workers at London hospital, The Lancet 395, 1608–1610 (2020).

28. C. J. Reynolds, L. Swadling, J. M. Gibbons, C. Pade, M. P. Jensen, M. O. Diniz, N. M. Schmidt, D. K. Butler, O. E. Amin, S. N. L. Bailey, S. M. Murray, F. P. Pieper, S. Taylor, J. Jones, M. Jones, W.-Y. J. Lee, J. Rosenheim, A. Chandran, G. Joy, C. D. Genova, N. Temperton, J. Lambourne, T. Cutino-Moguel, M. Andiapen, M. Fontana, A. Smit, A. Semper, B. O’Brien, B. Chain, T. Brooks, C. Manisty, T. Treibel, J. C. Moon, Covid. Investigators$, M. Noursadeghi, Covid. immune correlates Network$, D. M. Altmann, M. K. Maini, Á. McKnight, R. J. Boyton, Discordant neutralizing antibody and T cell responses in asymptomatic and mild SARS-CoV-2 infection, Science Immunology 5 (2020), doi:10.1126/sciimmunol.abf3698.

29. A. Roers, H. K. Hochkeppel, M. A. Horisberger, A. Hovanessian, O. Haller, MxA Gene Expression after Live Virus Vaccination: A Sensitive Marker for Endogenous Type I Interferon, J Infect Dis 169, 807–813 (1994).

30. J. Yu, D. R. Peterson, A. M. Baran, S. Bhattacharya, T. N. Wylie, A. R. Falsey, T. J. Mariani, G. A. Storch, Host Gene Expression in Nose and Blood for the Diagnosis of Viral Respiratory Infection, J Infect Dis 219, 1151– 1161 (2019)

31. E. Pairo-Castineira, S. Clohisey, L. Klaric, A. D. Bretherick, K. Rawlik, D. Pasko, S. Walker, N. Parkinson, M. H. Fourman, C. D. Russell, J. Furniss, A. Richmond, E. Gountouna, N. Wrobel, D. Harrison, B. Wang, Y. Wu, A. Meynert, F. Griffiths, W. Oosthuyzen, A. Kousathanas, L. Moutsianas, Z. Yang, R. Zhai, C. Zheng, G. Grimes, R. Beale, J. Millar, B. Shih, S. Keating, M. Zechner, C. Haley, D. J. Porteous, C. Hayward, J. Yang, J. Knight, C. Summers, M. Shankar-Hari, P. Klenerman, L. Turtle, A. Ho, S. C. Moore, C. Hinds, P. Horby, A. Nichol, D. Maslove, L. Ling, D. McAuley, H. Montgomery, T. Walsh, A. Pereira, A. Renieri, X. Shen, C. P. Ponting, A. Fawkes, A. Tenesa, M. Caulfield, R. Scott, K. Rowan, L. Murphy, P. J. M. Openshaw, Malcolm G. Semple, A. Law, V. Vitart, J. F. Wilson, J. K. Baillie, Genetic mechanisms of critical illness in Covid-19, Nature, 1–1 (2020).

32. Q. Zhang, P. Bastard, Z. Liu, J. L. Pen, M. Moncada-Velez, J. Chen, M. Ogishi, I. K. D. Sabli, S. Hodeib, C. Korol, J. Rosain, K. Bilguvar, J. Ye, A. Bolze, B. Bigio, R. Yang, A. A. Arias, Q. Zhou, Y. Zhang, F. Onodi, S. Korniotis, L. Karpf, Q. Philippot, M. Chbihi, L. Bonnet-Madin, K. Dorgham, N. Smith, W. M. Schneider, B. S. Razooky, H.-H. Hoffmann, E. Michailidis, L. Moens, J. E. Han, L. Lorenzo, L. Bizien, P. Meade, A.-L. Neehus, A. C. Ugurbil, A. Corneau, G. Kerner, P. Zhang, F. Rapaport, Y. Seeleuthner, J. Manry, C. Masson, Y. Schmitt, A. Schlüter, T. L. Voyer, T. Khan, J. Li, J. Fellay, L. Roussel, M. Shahrooei, M. F. Alosaimi, D. Mansouri, H. Al-Saud, F. Al-Mulla, F. Almourfi, S. Z. Al-Muhsen, F. Alsohime, S. A. Turki, R. Hasanato, D. van de Beek, A. Biondi, L. R. Bettini, M. D’Angio’, P. Bonfanti, L. Imberti, A. Sottini, S. Paghera, E. Quiros-Roldan, C. Rossi, A. J. Oler, M. F. Tompkins, C. Alba, I. Vandernoot, J.-C. Goffard, G. Smits, I. Migeotte, F. Haerynck, P. Soler-Palacin, A. Martin-Nalda, R. Colobran, P.-E. Morange, S. Keles, F. Çölkesen, T. Ozcelik, K. K. Yasar, S. Senoglu, Ş.N. Karabela, C. Rodríguez-Gallego, G. Novelli, S. Hraiech, Y. Tandjaoui-Lambiotte, X. Duval, C. Laouénan, C.-S. Clinicians†,C. Clinicians†, I.C. Group†, F.C.C.S. Group†, C.-C. Cohort†, A.U. C.-19 Biobank†, C.H.G. Effort†, N.-U.C.I. Group†, A. L. Snow, C. L. Dalgard, J. D. Milner, D. C. Vinh, T. H. Mogensen, N. Marr, A. N. Spaan, B. Boisson, S. Boisson-Dupuis, J. Bustamante, A. Puel, M. J. Ciancanelli, I. Meyts, T. Maniatis, V. Soumelis, A. Amara, M. Nussenzweig, A. García-Sastre, F. Krammer, A. Pujol, D. Duffy, R. P. Lifton, S.-Y. Zhang, G. Gorochov, V. Béziat, E. Jouanguy, V. Sancho-Shimizu, C. M. Rice, L. Abel, L. D. Notarangelo, A. Cobat, H. C. Su, J.-L. Casanova, Inborn errors of type I IFN immunity in patients with life-threatening COVID-19, Science 370 (2020), doi:10.1126/science.abd4570.

33. S. Rosebeck, D. W. Leaman, Mitochondrial localization and pro-apoptotic effects of the interferon-inducible protein ISG12a, Apoptosis 13, 562–572 (2008).

34. E. L. Tsalik, R. Henao, M. Aydin, C. Bullard, J. Montgomery, J. Nawrocki, M. Deneris, C. Gritzen, J. Jones, R. Crisp, M. T. Mcclain, T. Burke, G. S. Ginsburg, A. Hemmert, C. W. Woods, 2012. FilmArray® Measurement of Host Response Signatures RapidLy Discriminates Viral, Bacterial, and Non-infectious Etiologies of Illness, Open Forum Infect Dis 5, pS586 (2018).

35. R. R. Miller III, B. K. Lopansri, J. P. Burke, M. Levy, S. Opal, R. E. Rothman, F. R. D’Alessio, V. K. Sidhaye, N. R. Aggarwal, R. Balk, J. A. Greenberg, M. Yoder, G. Patel, E. Gilbert, M. Afshar, J. P. Parada, G. S. Martin, A. M. Esper, J. A. Kempker, M. Narasimhan, A. Tsegaye, S. Hahn, P. Mayo, T. van der Poll, M. J. Schultz, B. P. Scicluna, P. K. Klouwenberg, A. Rapisarda, T. A. Seldon, L. C. McHugh, T. D. Yager, S. Cermelli, D. Sampson, V. Rothwell, R. Newman, S. Bhide, B. A. Fox, J. T. Kirk, K. Navalkar, R. F. Davis, R. A. Brandon, R. B. Brandon, Validation of a Host Response Assay, SeptiCyte LAB, for Discriminating Sepsis from Systemic Inflammatory Response Syndrome in the ICU, American Journal of Respiratory and Critical Care Medicine 198 (2018), doi:10.1164/rccm.201712-2472OC.

36. R. K. Gupta, C. T. Turner, C. Venturini, H. Esmail, M. X. Rangaka, A. Copas, M. Lipman, I. Abubakar, M. Noursadeghi, Concise whole blood transcriptional signatures for incipient tuberculosis: a systematic review and patient-level pooled meta-analysis, The Lancet Respiratory Medicine 8, 395–406 (2020).

37. C. T. Turner, R. K. Gupta, E. Tsaliki, J. K. Roe, P. Mondal, G. R. Nyawo, Z. Palmer, R. F. Miller, B. W. Reeve, G. Theron, M. Noursadeghi, Blood transcriptional biomarkers for active pulmonary tuberculosis in a high-burden setting: a prospective, observational, diagnostic accuracy study, The Lancet Respiratory Medicine 8, 407–419 (2020).

38. N. L. Bray, H. Pimentel, P. Melsted, L. Pachter, Near-optimal probabilistic RNA-seq quantification, Nat Biotechnol 34, 525–527 (2016).

39. C. Soneson, M. I. Love, M. D. Robinson, Differential analyses for RNA-seq: transcript-level estimates improve gene-level inferences, F1000Research 4, 1521 (2016).

40. S. Durinck, Y. Moreau, A. Kasprzyk, S. Davis, B. De Moor, A. Brazma, W. Huber, BioMart and Bioconductor: a powerful link between biological databases and microarray data analysis, Bioinformatics 21, 3439–3440 (2005).

41. E. R. DeLong, D. M. DeLong, D.L. Clarke-Pearson, Comparing the areas under two or more correlated receiver operating characteristic curves: a nonparametric approach, Biometrics 44, 837–845 (1988).

42. A. K. Zaas, M. Chen, J. Varkey, T. Veldman, A. O. Hero, J. Lucas, Y. Huang, R. Turner, A. Gilbert, R. Lambkin-Williams,N.C. Øien, B. Nicholson, S. Kingsmore, L. Carin, C. W. Woods, G. S. Ginsburg, Gene Expression Signatures Diagnose Influenza and Other Symptomatic Respiratory Viral Infections in Humans, Cell Host & Microbe 6, 207–217 (2009).

43. O. Ramilo, W. Allman, W. Chung, A. Mejias, M. Ardura, C. Glaser, K. M. Wittkowski, B. Piqueras, J. Banchereau, A. K. Palucka, D. Chaussabel, Gene expression patterns in blood leukocytes discriminate patients with acute infections, Blood 109, 2066–2077 (2007).

44. R. Pescarmona, A. Belot, M. Villard, L. Besson, J. Lopez, I. Mosnier, A.-L. Mathieu, C. Lombard, L. Garnier, C. Frachette, T. Walzer, S. Viel, Comparison of RT-qPCR and Nanostring in the measurement of blood interferon response for the diagnosis of type I interferonopathies, Cytokine 113, 446–452 (2019).

45. Y. Huang, A. K. Zaas, A. Rao, N. Dobigeon, P. J. Woolf, T. Veldman, N.C. Øien, M. T. McClain, J. B. Varkey, B. Nicholson, L. Carin, S. Kingsmore, C. W. Woods, G. S. Ginsburg, A. O. Hero, Temporal dynamics of host molecular responses differentiate symptomatic and asymptomatic influenza A infection, PLoS Genet 7, e1002234 (2011).

