## Supplementary Materials for "Blood transcriptional biomarkers of acute viral infection for detection of pre-symptomatic SARS-CoV-2 infection"

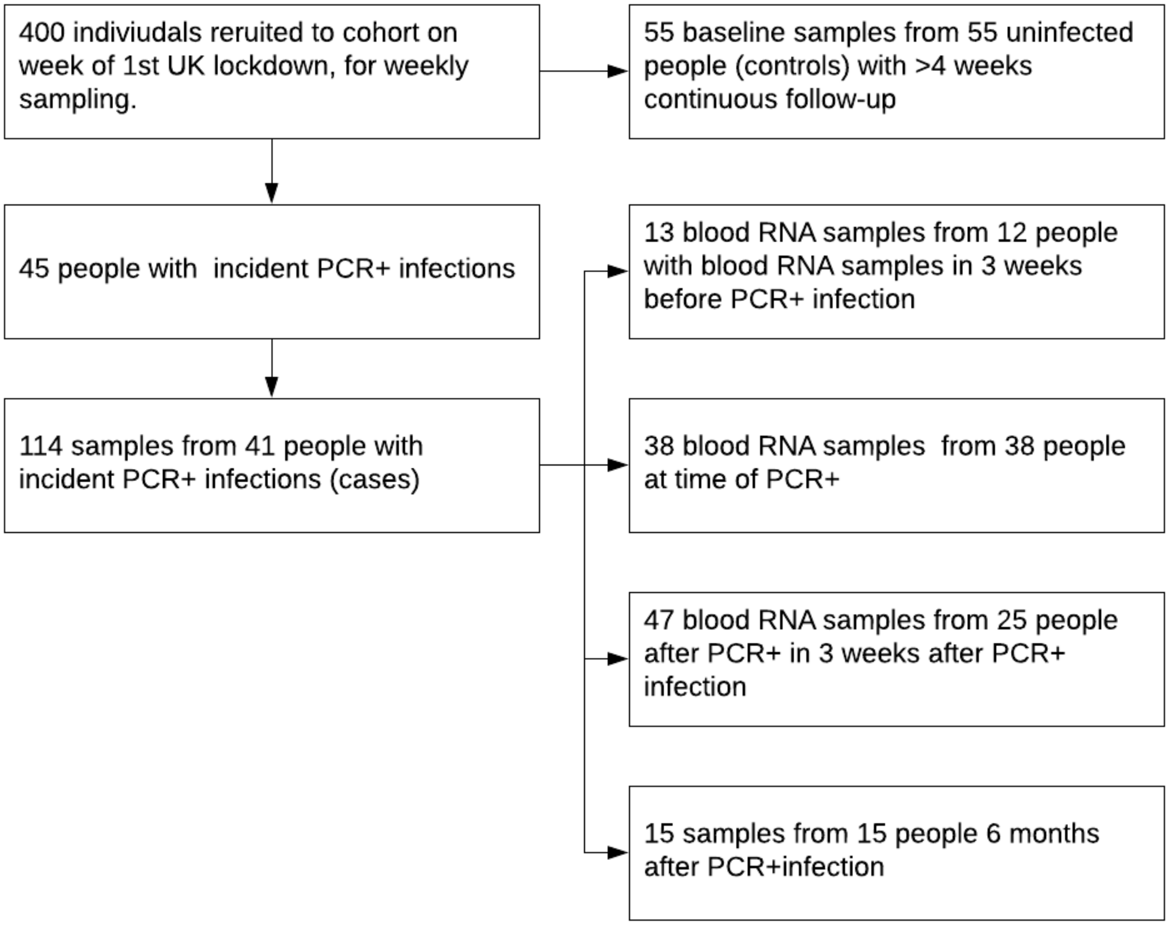


**Fig. S1. CONSORT flow diagram for samples from cases and controls included in this analysis.**

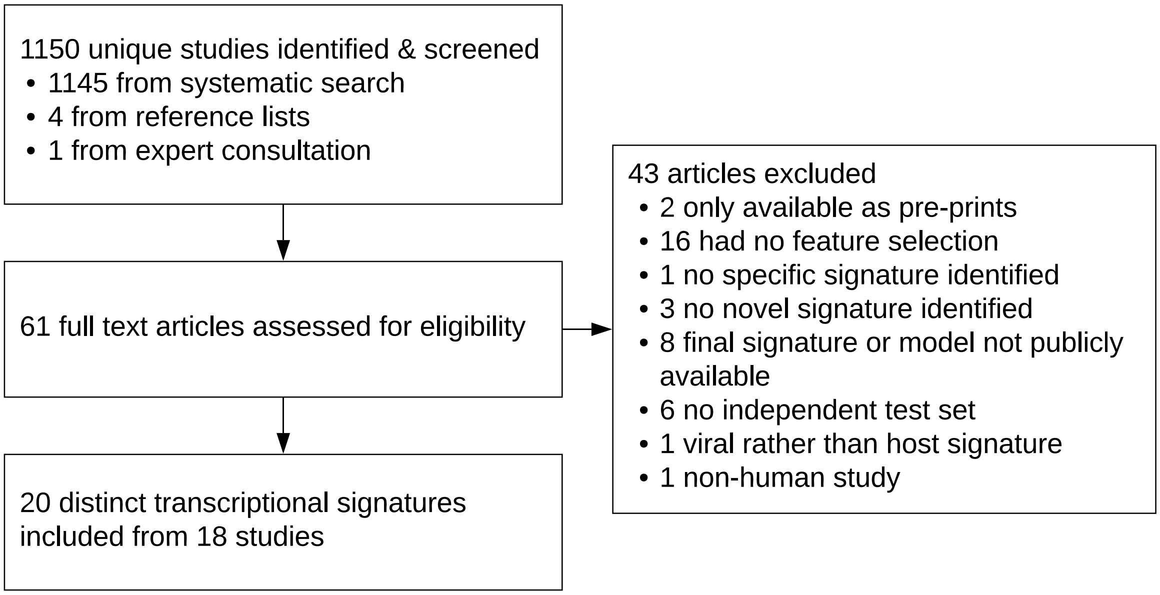


**Fig. S2. PRISMA flow-chart of systematic review process for identification of concise transcriptional signatures for the diagnosis of viral infections**

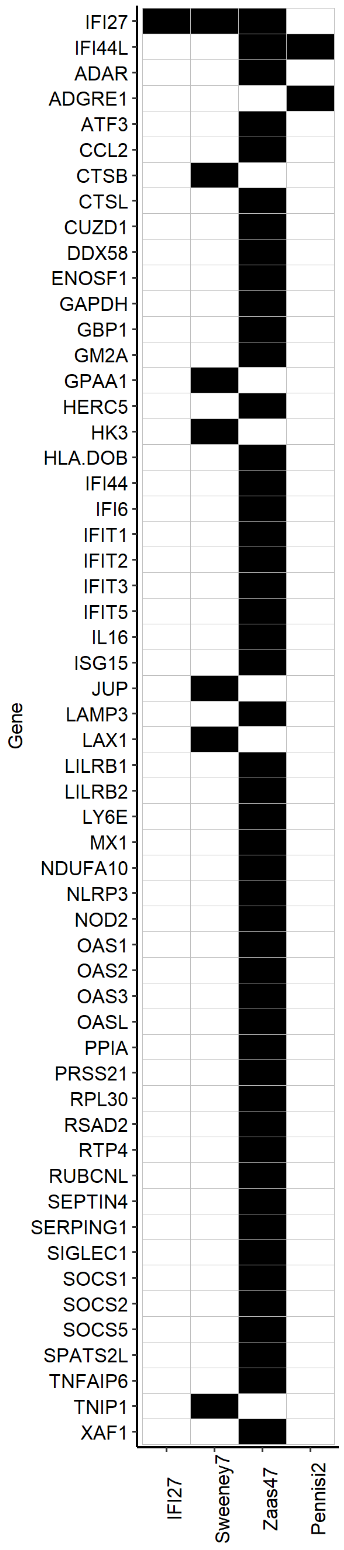


**Fig. S3. Constituent genes comprising four best performing RNA signatures for discriminating participants with contemporaneous PCR-confirmed SARS-CoV-2 infection, compared to uninfected controls**

| Characteristic | Overall, N = 96 | Cases, N = 41[1] | Controls, N = 55[1] |
|---|---|---|---|
| Age | 36 (27, 47) | 36 (28, 46) | 36 (26, 50) |
| Sex | | | |
| Female | 69 (72%) | 28 (68%) | 41 (75%) |
| Male | 27 (28%) | 13 (32%) | 14 (25%) |
| Ethnicity | | | |
| White | 66 (69%) | 25 (62%) | 41 (75%) |
| Black | 6 (6.3%) | 5 (12%) | 1 (1.8%) |
| Asian | 18 (19%) | 9 (22%) | 9 (16%) |
| Other | 5 (5.3%) | 1 (2.5%) | 4 (7.3%) |
| Unknown | 1 | 1 | 0 |
| Samples | | | |
| 1 | 67 (70%) | 12 (29%) | 55 (100%) |
| 2 | 7 (7.3%) | 7 (17%) | 0 (0%) |
| 3 | 6 (6.2%) | 6 (15%) | 0 (0%) |
| 4 | 10 (10%) | 10 (24%) | 0 (0%) |
| 5 | 6 (6.2%) | 6 (15%) | 0 (0%) |
| Case-defining symptoms | 36 (38%) | 31 (76%) | 5 (9.1%) |

[1]Statistics presented: median (IQR); n (%)

**Table S1. Baseline characteristics of the study cohort.**

| Signature | AUROC | Sensitivity | Specificity |
|---|---|---|---|
| IFI27 | 0.96 (0.92 - 1) | 0.86 (0.69 - 0.95) | 0.95 (0.85 - 0.98) |
| Sweeney7 | 0.95 (0.9 - 1) | 0.86 (0.69 - 0.95) | 0.95 (0.85 - 0.98) |
| Zaas48 | 0.94 (0.89 - 0.99) | 0.62 (0.44 - 0.77) | 0.95 (0.85 - 0.98) |
| Pennisi2 | 0.92 (0.86 - 0.97) | 0.62 (0.44 - 0.77) | 0.95 (0.85 - 0.98) |
| IFI44L | 0.91 (0.85 - 0.97) | 0.55 (0.38 - 0.72) | 0.95 (0.85 - 0.98) |
| AndresTerre11 | 0.9 (0.83 - 0.96) | 0.55 (0.38 - 0.72) | 0.95 (0.85 - 0.98) |
| Henrickson16 | 0.89 (0.82 - 0.97) | 0.55 (0.38 - 0.72) | 0.93 (0.83 - 0.97) |
| Lydon15 | 0.88 (0.81 - 0.96) | 0.59 (0.41 - 0.74) | 0.95 (0.85 - 0.98) |
| TrouilletAssant6 | 0.88 (0.8 - 0.95) | 0.52 (0.34 - 0.69) | 0.93 (0.83 - 0.97) |
| Herberg2 | 0.85 (0.77 - 0.94) | 0.48 (0.31 - 0.66) | 0.93 (0.83 - 0.97) |
| RSAD2 | 0.84 (0.75 - 0.93) | 0.48 (0.31 - 0.66) | 0.93 (0.83 - 0.97) |
| Sampson4 | 0.84 (0.75 - 0.93) | 0.52 (0.34 - 0.69) | 0.93 (0.83 - 0.97) |
| MX1 | 0.83 (0.73 - 0.92) | 0.45 (0.28 - 0.62) | 0.95 (0.85 - 0.98) |
| Sampson10 | 0.82 (0.72 - 0.92) | 0.52 (0.34 - 0.69) | 0.95 (0.85 - 0.98) |
| Tsalik33 | 0.8 (0.7 - 0.9) | 0.41 (0.26 - 0.59) | 0.98 (0.9 - 1) |
| Lopez7 | 0.79 (0.69 - 0.9) | 0.38 (0.23 - 0.56) | 0.98 (0.9 - 1) |
| IFIT3 | 0.76 (0.64 - 0.88) | 0.45 (0.28 - 0.62) | 0.93 (0.83 - 0.97) |
| OLFM4 | 0.6 (0.47 - 0.73) | 0 (0 - 0.12) | 0.98 (0.9 - 1) |
| Sweeney11 | 0.57 (0.43 - 0.71) | 0.1 (0.04 - 0.26) | 0.96 (0.88 - 0.99) |
| Yu3 | 0.57 (0.43 - 0.71) | 0.07 (0.02 - 0.22) | 1 (0.93 - 1) |

**Table S2. Performance metrics of whole-blood RNA signatures for discrimination of participants with PCR-confirmed SARS-CoV-2 infection at week of first week of PCR-positivity (PCR+ve_0) from uninfected controls, excluding participants with contemporaneous symptoms at the time of nasopharyngeal swab sampling.** Discrimination is shown as area under the receiver operating characteristic curve (AUROC). Sensitivity and specificity are shown using pre-defined thresholds of 2 standard deviations above the mean of the uninfected control population (Z2). All metrics as shown as point estimates (95% confidence intervals).

| Signature | AUROC | Sensitivity | Specificity |
|---|---|---|---|
| Henrickson16 | 0.83 (0.71 - 0.96) | 0.3 (0.11 - 0.6) | 0.93 (0.83 - 0.97) |
| IFIT3 | 0.82 (0.69 - 0.94) | 0.3 (0.11 - 0.6) | 0.93 (0.83 - 0.97) |
| AndresTerre11 | 0.82 (0.65 - 0.99) | 0.3 (0.11 - 0.6) | 0.95 (0.85 - 0.98) |
| Pennisi2 | 0.8 (0.64 - 0.95) | 0.3 (0.11 - 0.6) | 0.95 (0.85 - 0.98) |
| Zaas48 | 0.78 (0.6 - 0.97) | 0.4 (0.17 - 0.69) | 0.95 (0.85 - 0.98) |
| TrouilletAssant6 | 0.78 (0.6 - 0.96) | 0.3 (0.11 - 0.6) | 0.93 (0.83 - 0.97) |
| IFI27 | 0.78 (0.59 - 0.97) | 0.4 (0.17 - 0.69) | 0.95 (0.85 - 0.98) |
| IFI44L | 0.77 (0.6 - 0.95) | 0.3 (0.11 - 0.6) | 0.95 (0.85 - 0.98) |
| RSAD2 | 0.77 (0.6 - 0.95) | 0.3 (0.11 - 0.6) | 0.93 (0.83 - 0.97) |
| Sampson4 | 0.75 (0.58 - 0.93) | 0.3 (0.11 - 0.6) | 0.93 (0.83 - 0.97) |
| Sweeney7 | 0.75 (0.52 - 0.98) | 0.4 (0.17 - 0.69) | 0.95 (0.85 - 0.98) |
| Herberg2 | 0.73 (0.54 - 0.92) | 0.3 (0.11 - 0.6) | 0.93 (0.83 - 0.97) |
| Lydon15 | 0.7 (0.49 - 0.91) | 0.3 (0.11 - 0.6) | 0.95 (0.85 - 0.98) |
| Tsalik33 | 0.68 (0.5 - 0.86) | 0.2 (0.06 - 0.51) | 0.98 (0.9 - 1) |
| MX1 | 0.67 (0.43 - 0.92) | 0.3 (0.11 - 0.6) | 0.95 (0.85 - 0.98) |
| Yu3 | 0.63 (0.44 - 0.83) | 0 (0 - 0.28) | 1 (0.93 - 1) |
| Lopez7 | 0.62 (0.39 - 0.85) | 0.2 (0.06 - 0.51) | 0.98 (0.9 - 1) |
| Sampson10 | 0.57 (0.32 - 0.83) | 0.3 (0.11 - 0.6) | 0.95 (0.85 - 0.98) |
| OLFM4 | 0.49 (0.29 - 0.69) | 1 (0.72 - 1) | 0.02 (0 - 0.1) |
| Sweeney11 | 0.49 (0.26 - 0.72) | 0.9 (0.6 - 0.99) | 0.04 (0.01 - 0.12) |

**Table S3. Performance metrics of whole-blood RNA signatures whole-blood RNA signatures for discrimination of participants with PCR-confirmed SARS-CoV-2 infection at week prior to first week of PCR-positivity (PCR+ve_-1).** Discrimination is shown as area under the receiver operating characteristic curve (AUROC). Sensitivity and specificity are shown using pre-defined thresholds of 2 standard deviations above the mean of the uninfected control population (Z2). All metrics as shown as point estimates (95% confidence intervals).

| |
|---|
| 1. Biomarkers/ |
| 2. biomarker*.tw. |
| 3. diagnostic*.tw. |
| 4. signature*.tw. |
| 5. classifier*.tw. |
| 6. 1 or 2 or 3 or 4 or 5 |
| 7. Viruses/ |
| 8. viral.tw. |
| 9. 7 or 8 |
| 10. RNA/ |
| 11. Transcriptome/ |
| 12. (transcriptom* or transcriptional).tw. |
| 13. (rna or mrna).tw. |
| 14. gene expression.tw. |
| 15. 10 or 11 or 12 or 13 or 14 |
| 16. Blood/ |
| 17. (host or blood).tw. |
| 18. 16 or 17 |
| 19. 6 and 9 and 15 and 18 |
| 20. remove duplicates from 19 |
| 21. limit 20 to "humans only (removes records about animals)" |

**Table S4. Search strategy for systematic Medline search, performed on 12/10/2020.**

Hakam Abbass, Aderonke Abiodun, Mashael Alfarih, Zoe Alldis, Daniel M Altmann, Mervyn Andiapen, Jessica Artico, Joao Augusto, Georgina L Baca, Anish Bhuva, Alex Boulter, Ruth Bowles , Rosemary J Boyton, Olivia Bracken, Timothy Brooks, Natalie Bullock, Gabriella Captur, Benny Chain, Nicola Champion, Carmen Chan, Jorge Couto de Sousa, Xose Couto-Parada , Marie-Teresa Cutino-Moguel, Rhodri H Davies, Keenan Dieobi-Anene, Karen Feehan, Malcolm Finlay, Marianna Fontana, Nasim Forooghi, Joseph M Gibbons, Derek Gilroy, Peter Griffiths, Rishi K Gupta, Matt Hamblin, Lauren M Hickling, Aroon D Hingorani, Lee Howes, Ivie Itua, Victor Jardim, Melanie Jensen, Meleri Jones, George Joy, Vikas Kapil, Jonathan Lambourne, WY Jason Lee, Mala K Maini, Vineela Mandadapu, Charlotte Manisty, Aine McKnight, Katia Menacho Medina, Celina Mfuko, Oliver Mitchelmore, James C Moon, Mahdad Noursadeghi, Ben O'Brien , Ben Ollivere, Corinna Pade, Susana Palma, Kush Patel, Ruth Parker, Brian Piniera, Alicja Rapala, Amy Richards, Mathew Robathan, Genine Sambile, Amanda Semper, Andreas Seraphim, Angelique Smit, Michelle Sugimoto, George D Thornton, Thomas A. Treibel, Arthur Tucker, Ana Valdes, Jessry Veerapen, Mohit Vijayakumar, Timothy Warner, Sophie Welch, Dylan Williams, Theresa Wodehouse , Lucinda Wynne, and Dan Zahedi.

**Table S5. COVIDsortium investigators (alphabetical order).**
